# Status of water sources, hygiene and sanitation and its impact on the health of households of Makwane Village, Limpopo Province, South Africa

**DOI:** 10.1101/2020.01.31.20019679

**Authors:** P Budeli, RC Moropeng, L Mpenyana-Monyatsi, I Kamika, MNB Momba

**Author notes:** To whom all correspondence should be addressed.

## Abstract

The key to reducing or even eradicating the burden of waterborne diseases is through appropriate sanitation facilities and piped water systems. Installation of centralised system may take decades to be established, especially in impoverished rural communities of African countries. A survey of 88 households representing Makwane, a scattered settlement in South Africa, Limpopo Province, was conducted to assess the status of basic services. A questionnaire was designed to obtain the required information, such as improved water sources, improved sanitation facilities, hygiene practices and incidence of diarrhoeal diseases in the community. A house-to-house survey was conducted from July to August in 2014 and data were collected from the heads of each household. Results of the survey revealed a complete absence of improved drinking water sources in the community (100%). People rely on any available water sources such stream water, or on rainwater harvesting. Safe hygiene practices were observed in most households with regards to water storage as they store water in 25 L plastic buckets (57%), vessels stored inside a room (76%), use storage containers covered with a lid (76%) and wash these containers at any time prior to storing water (39%). Results also indicated a high percentage of households not treating water (81%) prior to use, disposing wastewater in the yard (97%), lacking access to improved sanitation facilities (41%), and not allowing children under 12 years old to use the toilets (62%). As a result, they practise open defecation as an alternative sanitation facility (86%). The main water source for the community was found to be the stream (31%) and this source is used for adequate personal hygiene in terms of full body bath (94%). In terms of health outcomes, the most prevalent health problem was found to be diarrhoea (75%), which occurred mostly in children less than 5 years old and was found to persist up to 3 days (34%). The community generally visited the clinic (75%) in cases of health problems. The implementation of point-of-use household drinking water treatment in Makwane households for the production of safe drinking water is highly recommended. In addition to this, a special education with emphasis on drinking water storage, cleaning of water storage containers and safe disposal of wastewater should be offered. Open defecation should also be discouraged to mitigate the bacterial contamination of water sources and transmission of diseases.

## 1. Introduction

Access to safe drinking water sources, appropriate sanitation facilities and good hygiene are not only fundamental to health and survival of the people, but also to the economic growth and the development of a country. These basic necessities are still a luxury for many people in the developing world especially in rural areas (Peter, 2010). Sustainable development goal 6 stipulates that we need to invest in adequate infrastructure, provide sanitation facilities, and encourage hygiene at every level in order to ensure universal access to safe and affordable drinking water for all by 2030. In addition to this, water related ecosystems such as forests, mountains, wetlands and rivers have to be protected to mitigate water scarcity. More international cooperation is also needed to encourage water efficiency and support treatment technologies in developing countries (UNDP, 2015). An estimate of 768 million people relied on ‘unimproved’ water supplies (as defined by the WHO/UNICEF Joint Monitoring Program for Water and Sanitation – JMP) which are thought to harbour high concentration of pathogenic contaminants and also more than 2.5 billion people are still deprived of access to an improved sanitation facility with 946 million people practising open defecation (WHO/UNICEF, 2013). Numerous water sources are classified as improved but are still not safe for human consumption (Bain et al., 2014). Improved sanitation includes flush or pour-flush toilet/latrine connected to a piped sewer system, a septic tank or a pit latrine, ventilated improved pit latrine, pit latrine with slab, or composting toilet (WHO/UNICEF JMP, 2010).

In low and middle-income countries, inadequate drinking water, sanitation, and hygiene have adverse effects in non-household settings, such as schools, health care facilities, and workplaces; this in turn affects the health, education, welfare, and productivity of populations. Inadequate hand hygiene practices have been estimated to affect 80% of the population globally (Freeman et al., 2014). A study by (Adukia, 2013) revealed that a lack of gender-separated toilets at schools negatively affects attendance of girls. Disabled persons face physical and social challenges related to inadequate drinking water, sanitation, and hygiene, which prevent them from attending schools, gaining employment, and using public services and amenities (Groce et al., 2011). Improper management of human excreta from sick patients in health care facilities poses threat to the health of the public to in the nearby communities. Transmission of infectious disease in non-household settings is most likely to cause larger epidemics as compared to household settings (Cronk et al., 2015).

Diarrhoea alone has been reported to be the third largest cause of morbidity and the sixth largest cause of mortality (Pond et al., 2004). While cholera mortality cases have been reported from all regions of the world, it has been proven that the poor socio-economic conditions and lack of access to improved drinking water sources and sanitation have led the African continent to be a major contributor of this waterborne disease. The recent estimate indicates 2.9 million of cases and 95,000 deaths in 69 endemic countries, with the majority of the burden reported in Sub-Saharan Africa (Ali et al., 2015). South Africa’s population stands at approximately 51.77 million, of whom 40% reside in rural areas. In spite of South African Government’s effort to provide safe drinking water and improved sanitation to all, huge challenges still remain in rural areas. While access to safe drinking for indigent households has increased to 71.6% in 2011, sanitation and sewerage services were only up to 57.9% (Statistics South Africa, 2013). This report also indicates that there was a decrease in the incidence of diarrhoea per 1 000 children under five years, down from 121.4 per 1 000 children in 2004 to 102.1 children in 2011.

An in-depth understanding of rural drinking water sources, sanitation and hygiene challenges is crucial in protecting the public health, improving the quality of life for rural residents and in developing a strong local economy (Phaswana-Mafuya, 2006a; 2006b). Against this background, the current study was conducted to determine the status of water sources, hygiene and sanitation and its impact on the health of household members of the Makwane Village, Limpopo, South Africa. The households in this community are scattered and some are several kilometres away from each other. The current study aims to generate information on the health status of rural communities and also provides critical information which policy makers may consider in order to meet the basic human needs of this community.

## 2. Materials and methods

### 2.1 Study area and population

This study was conducted in a rural village called Makwane, located near Roossenekal, which is under the jurisdiction of Elias Motsoaledi Municipality, Sekhukhune District, in Limpopo Province, South Africa. Makwane is one of the remote villages in this district, facing severe problems linked to lack of access to safe water and adequate sanitation facilities. Springs and streams are the main water sources of the village of Makwane. The majority of people residing in the Makwane village speak Sepedi. Makwane Village is currently divided into four sections namely: (a) Lepururu, with 21 households; (b) Ditakaneng, with 23 households; (c) new stands with 39 households; and (d) Nkakaboleng with 11 households see also (S1 Map). The village had a total population size of 480 at the time of the study and 94 households.

### 2.2 Data collection

For the purpose of this study, a survey questionnaire was developed to gather general information on the following; (i) socio-economic status of household members, which includes the age and gender of people dwelling in households. (ii) type of water delivery system used which includes: type of water source, frequency of water collection per day by household, distance from the collection point to the household, type of water treatment used prior to use; (iii) type of sanitation practices; and (iv) health and hygiene practices, see also (S2 file). The questionnaire was also translated into the local language in order for the Sepedi speaking participants to understand.

The communities were visited from Monday to Friday between 11:00am to 05:00pm. All the 94 households were approached to participate and 88 agreed to be interviewed (94%). Therefore, information leaflets and informed consent forms were distributed to and signed by all 88 households that agreed to participate in the study see also (S3 file). The study made use of face-to-face interviews where people were interviewed in their homes. The information about hygiene and sanitation issues in every household was collected in a single session. The interviewers administered structured questionnaires to the heads of each household who were aware of the hygienic conditions in the household. In cases where the heads were not aware of hygiene conditions in the household, women were involved because they are the main caregivers in homebased duties like fetching water and maintaining hygiene. In terms of prevalence and incidence of waterborne and hygiene-related diseases in households across the Makwane Village, the main focus was diarrhoea although other diseases were also considered. These included trachoma, body lice and hot tub rash. The diarrhoea diary was used to record the incidence of diarrhoea and the duration of diarrhoea in members dwelling in each household. The interview was estimated to last approximately 60 minutes on average.

## 3. Results

### 3.1 Population demographics

The population demographics of Makwane Village are shown in Figure 1. About 51% of the population were males and 49% of the population were females. The majority of residents were aged between 22-55 years old while the smallest group was identified to be above 55 years old.

**Figure 1:**
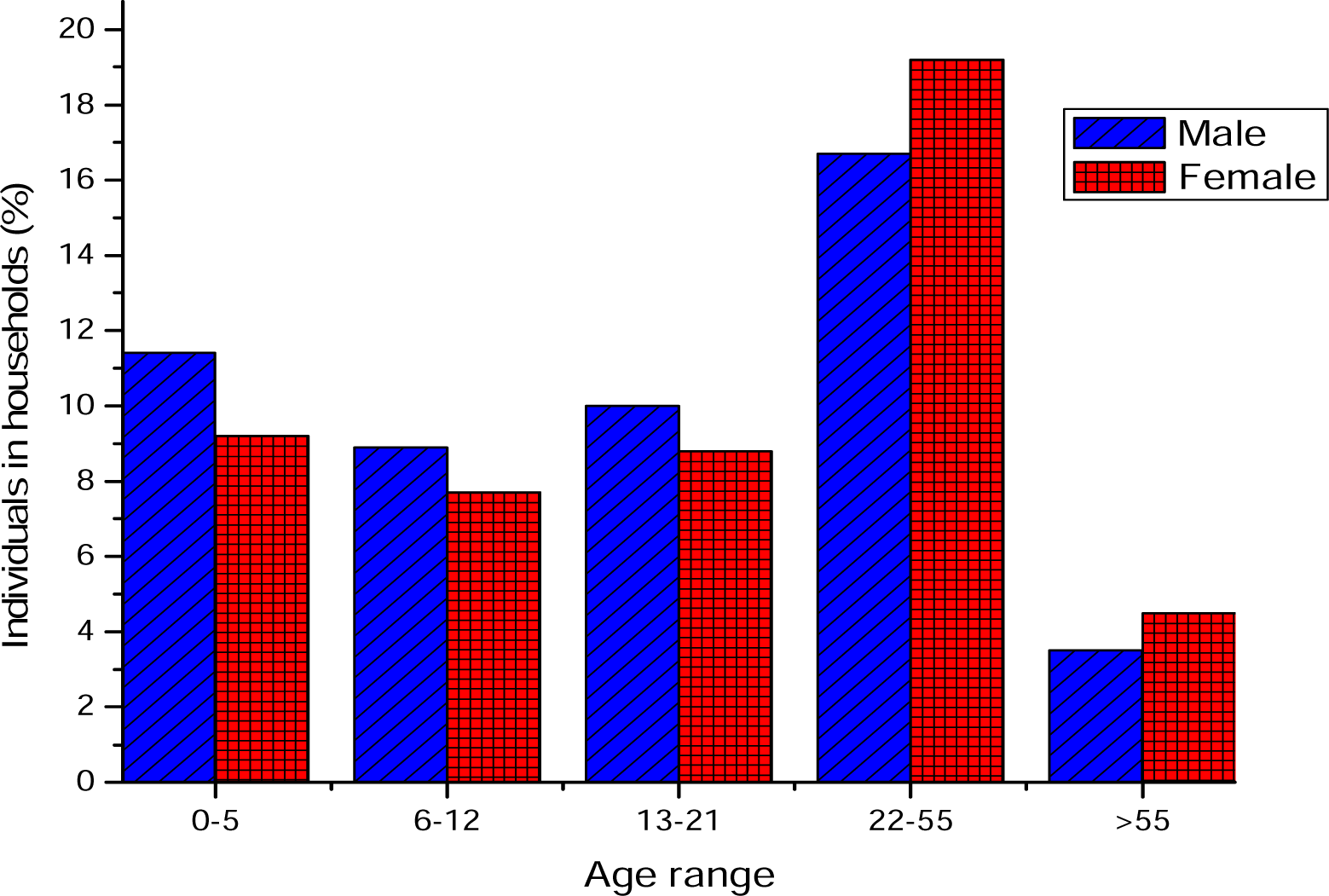
Population demographics of Makwane Village.

### 3.2 Source of water

The findings of the current study show that, almost three-quarters of the community (70%) generally depend on the stream as their source of water for domestic use and other purposes though the quality and quantity of the water source may vary depending on the seasons (dry or wet season). The majority of households (56%) collect water more than three times a day from the water source. Of all the households, 47% collect water once a day from the stream while 58% could collect at least three buckets in one visit from the stream. Furthermore, 57% of household members do not walk further than 100 m to collect water, as they are located closer to the communal standpipe. Roof harvesting of rainwater is used only during wet seasons (7%) and 45% of the population use spring water. The community uses two types of containers: plastic buckets and narrow mouth plastic jerry cans. During the study, a plastic bucket with a lid was found to be the most frequently used container (52%) for the collection of water, while 30% of households used plastic bucket without a lid. Plastic jerry can without lid was found to be the type of container least often used by the Makwane community. In terms of protection of water sources, the entire community confirmed that the main water source is not protected, although there are no agricultural practices along the water source.

### 3.3 Treatment and water storage in the households

The findings revealed that almost the entire community (81%) used the collected water for drinking or cooking purposes without any prior treatments. Furthermore, only 11% of the population used liquid bleach prior to usage, followed by 8% who boiled water before usage. The community uses four types of storage containers: plastic bucket, metallic bucket, plastic jerry can and plastic drum. The findings revealed that water was generally stored in 25 L plastic buckets (59%), and the buckets were generally kept inside the room (90%). In terms of protection of the storage containers, 78% of households stored their containers fully covered, 3% half covered them and 18% do not cover their containers completely. Almost two-thirds of the households in Makwane Village used the same container for both collection and storage and 38% used different containers. Less than (40%) of the households washed their containers every time they collected and stored water, although other households washed their containers once a day (25%), once a week (22%) and once a month (13%). Generally, the disinfectants were not used to wash the storage containers (86%). It was found that all households (100%) immersed their household utensils to withdraw water from the storage containers.

### 3.4 Hygiene status in the households and trend of sanitation facility

Figure 2 illustrates the hygiene practices and the source of water used to maintain the hygienic conditions in each household across the village. During the study, the majority of the householders practiced safe personal hygiene as they indulged in regular body bath (94%). Approximately half (51%) of the population used untreated water for bathing. An additional hygiene challenge noted within the community was the fact that children under the age of 12 in most of the households (62%) were not allowed to use the toilet. The reason attributed to this practice was lack of improved sanitation facilities; parents feared that their children are at risk of falling into and drowning in outside toilets, such as pit latrines.

**Figure 2:**
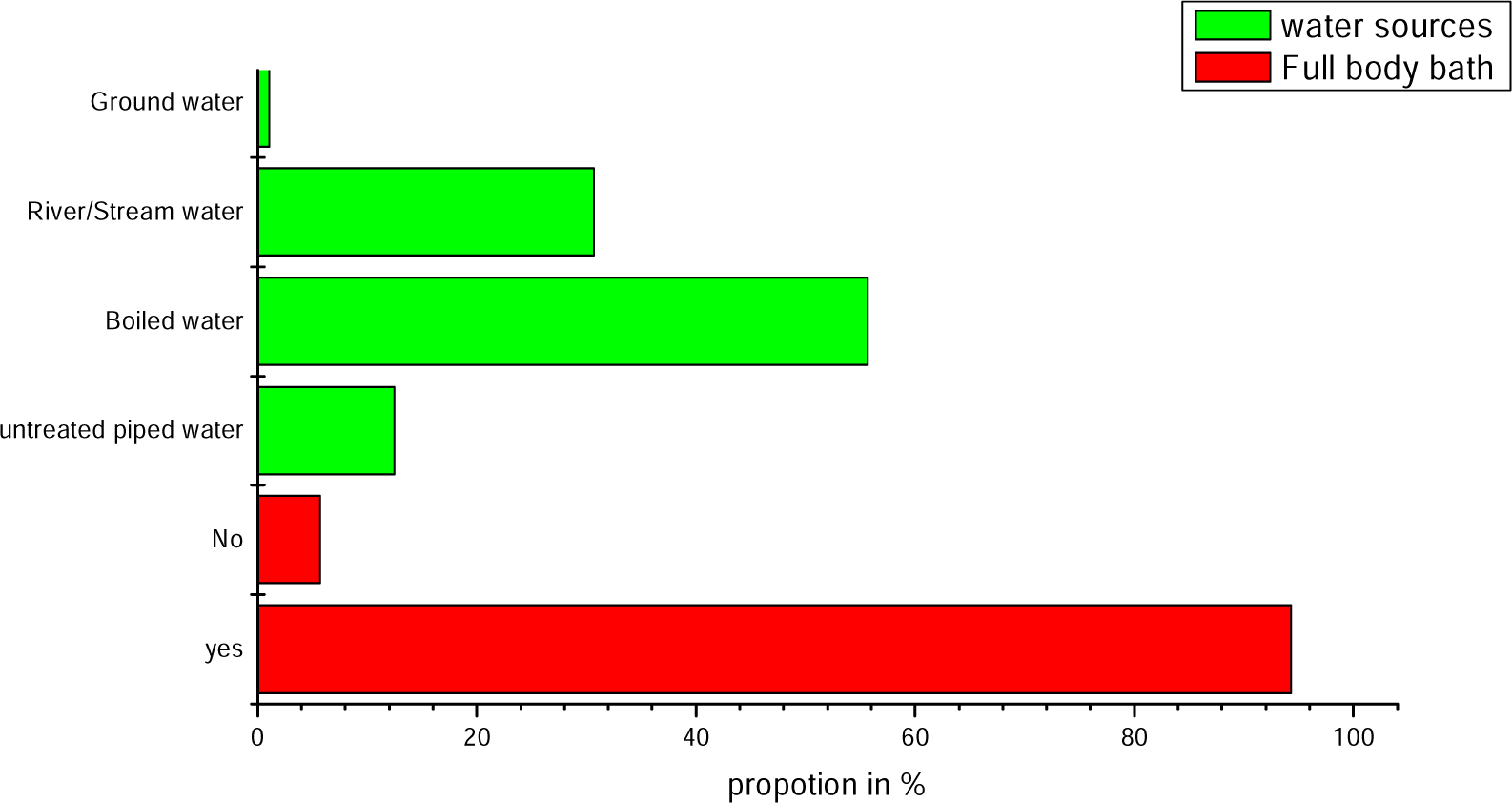
Results pertaining to hygiene status and sources of water in the householdss.

Besides the hygiene challenges, major sanitation issues were observed across the village. Approximately 41% of the households in the community did not have toilets installed in their yards and 86% of the households use the open field as an alternative for sanitation facility while the remaining 13% share sanitation facilities with their neighbours. The findings showed that 60% of householders safely dispose their solid waste (e.g. diapers) in the pit latrines, while 6% dispose of soiled diapers in the streams, 9% in the field, 37% in a separate pit and 11% burn them. Only 25% of the households use reusable diapers.. Households using reusable diapers generally dispose of the faecal matter by soaking the soiled fabric diapers in water and then discarding the wastewater in an open field. While it is encouraging to note that most of the households safely dispose their waste, a sizeable percentage of the households where unsafe disposal is practised should also be taken into consideration. As for the disposal of wastewater generated from various domestic uses, it was found that approximately 97% of the households discard their wastewater in the field, while 2% use it for gardening and 1% for construction purposes.

### 3.5 Health-related incidents, occurrence of diarrhoea in the households and residents attending primary health care facilities

The prevalence of diseases occurring in households in descending order was as follows: diarrhoea (75%), hot tub rash (17%), body lice (3%) and trachoma (2%). These figures show that diarrhoeal diseases affect (75%) of the households although some households experienced two or three of the diseases mentioned above from time to time. Moreover, 15% of all the households surveyed during this study did not feel comfortable disclosing the information.. Regardless of the health-related incidents within the community, 75% of the households regularly visited the community health care facility and almost 20% of households did not visit any health care facility in case of health-related incidents. Failure to take a sick child to a clinic may lead to serious complications especially in children under the age of 5 years due to a weak immune system. It is assumed that the community is reluctant to go to the clinic due to financial constraints and distance as the clinic is located far from the village.

## 4. Discussion

The present study assessed the status of basic services in 88 households and revealed that there is a high number of young people, especially children under the age of 5 years old (21%) indicated the potential vulnerability of this community in case of any issues related to access to safe drinking water, improved sanitation facilities and hygiene practices. The present study further revealed that Makwane Village depends mostly on a stream (70%) as the most reliable water source although this water source is not protected from contamination of any kind. In order to address this problem, one of the approaches that can be used is the installation of protected sources such as boreholes, standpipes or wells to provide water of improved quality. In Makwane Village, water storage is a common practice, with 59% of the population storing water using 25 L plastic buckets, highlighting the absence or inefficiency of communal standpipes. Furthermore, more than half of the households in the community use the same container for both collection and storage. Although most of the households are practising safe water storage, it is equally imperative that the number of the households who are not practising safe water storage should not be overlooked as inappropriate storage of water plays an important role in the deterioration of its quality. The present results also show that 86% households did not use disinfectants to clean the storage containers. This indicates the severity of the issue as approximately 20% of the population is children under the age of 5 years old (Figure 1). Since unsafe storage constitute a public threat as it can lead to water contamination, Safe water storage and handling practices should be promoted from households. However, in the current study, practices of immersing household utensils into storage vessels to withdraw water and touching of taps are widespread (100%) and this practice has been reported to be a route for the transfer of faecal contaminants (Quick et al.,1999). In addition to the untreated sources of water and inappropriate storage of the water, the present findings revealed that households (81%) across Makwane Village do not treat their water before drinking. The practice of drinking water without any prior treatment has been associated with health effects such as gastrointestinal and stomach illnesses like nausea, vomiting, cramps, and diarrhoea (WHO/UNICEF, 2010).

In contrast, this study further revealed that most of the householders in the community (94%) practise good personal hygiene by regular full body bathing, even though 30% used water from a stream as the sole water source for the above purpose. Other hygiene related illness that are attributed to irregular body bath includes but not limited to trachoma, body lice, etc. This study also showed that children up to the age of 12 years old were encouraged to do open defecation even when toilet facilities were present. The reason for this could be attributed to the poor standard and infrastructure of the toilets it was estimated that at least 20 people die each year in South Africa (KwaZulu-Natal Province) as a result of falling into the pit poor standard latrines and drowning (Personal communication, 1993). It should be mentioned that lack of access to sanitation facilities does not only affect the health of individuals but it also compromises the individual’s right to human dignity. The most recent StatsSA census has documented that two-thirds of Limpopo Province residents still do not have access to sufficient sanitation (Statistics South Africa, 2011). This was also confirmed in the current results as 97% of households are reported to discard wastewater in their yards or immediate environment as there is no drainage system. Based on these findings, there is a need to educate the householders about waste management, especially in this area where access and supply of safe drinking water is still a dream. Greywater disposed of in the yard as opposed to the vegetable patch also plays a role in the transmission of pathogens such as mosquito flies. In general, the incidence of diarrhoea occurred more frequently in 38% of the households with the highest prevalence occurring among children ≤5 years (74%) and was found to persist up to ≥ 3 days in most children (45%). As diarrhoea was the most prevalent health incidence among the Makwane households, a further investigation was done to ascertain the frequency and type of diarrhoea among the affected households. This study suggests that the burden of waterborne and other hygiene related diseases cannot be mitigated by provision of safe drinking water as a stand-alone intervention, but rather by adopting a holistic approach that includes good hygiene practices, improved sanitation facilities and access to safe drinking water sources.

## 5. Conclusion and recommendations

In general, unsafe hygiene practices were observed in the Makwane community, in terms of water source protection, sanitation and water storage, and the high prevalence of diarrhoea and other waterborne and hygiene-related diseases in this community could be attributed to these practices. Further studies are needed to investigate and assess the general water quality of various sources used for drinking-water purposes in this area so as to ascertain the health risk that may be posed by these water sources. This study highly recommends the implementation of point-of-use household drinking-water treatment in Makwane households for the production of safe drinking water coupled with a community-based hygiene education focusing on drinking water storage containers, open defecation and safe disposal of wastewater.

## Data Availability

We have included all the data referred to in this manuscript

## Ethical Approval

The study was conducted in accordance with the Declaration of Helsinki, and approved by the Faculty of Science Research Ethics Committee (FCRE) at the Tshwane University of Technology (TUT), where the study was registered Ref: FCRE 2014/07/20. Furthermore, the authorization to conduct the study in Makwane Village was obtained from Elias Motswaledi Municipality’s Manager, the Municipal Councillor and the Committee. The informed consent was obtained from the participants of the selected households prior to commencement of the project

## Acknowledgement

We wish to express our appreciation to the National Research Foundation (NRF) of South Africa for funding this research and Tshwane University of Technology (TUT) for the top-up scholarship awarded to the student.

## Conflict of interest

The authors declare that there are no conflicts of interest.

## Notes

### Competing Interest Statement

The authors have declared no competing interest.

## References

Adukia, A., 2013. Sanitation and Education. Harvard University, Cambridge, Retrieved from: http://scholar.harvard.edu/files/adukia/files/adukia sanitation and education.pdf (07.09.14).

Alcock, P.G., 1999. A water resources and sanitation systems source book with special reference to Kwa-Zulu Natal. Part 6. WRC Report No. 384/6/99. Water Research Commission, Pretoria, South Africa.

Ali, M., Nelson, A.R., Lopez, A.L., Sack, D.A., 2015. Updated global burden of cholera in endemic countries. PLoS neglected tropical diseases, 9(6), p.e0003832.

Allegranzi, B., Nejad, S.B., Combescure, C., Graafmans, W., Attar, H., Donaldson, L., Pittet, D., 2011. Burden of endemic health-care-associated infection in developing countries: systematic review and meta-analysis. Lancet 377 (9761), 228–241.

Bain, R., Cronk, R., Hossain, R., Bonjour, S., Onda, K., Wright, J., Yang, H., Slaymaker, T., Hunter, P., Prüss-Ustün, A., Bartram, J., 2014. Global assessment of exposure to faecal contamination through drinking water based on a systematic review. Tropical Medicine & International Health, 19(8), 917–927.

Chidavaenzi, M.T., Jere M., Nhandara C., Chingundury, D., Bradley, M., 1998. An evaluation of water urns to maintain domestic water quality, in: Pickford, J. (Ed.) Proceedings of the 24th WEDC Conference, Islamabad, Pakistan. WEDC, Loughborough, pp. 249–253.

Cronk, P., Slaymaker, T., Bartram, J., 2015.Monitoring drinking water, sanitation, and hygiene in non-household settings: Priorities for policy and practice. International Journal of Hygiene and Environmental Health 218, 694–703.

Department of National Health and Population Development, 2001. National Status Report on Cholera Epidemic in South Africa. Available at: http://sandmc.pwv.gov.za/ndmc/cholera/.

Freeman, M.C., Clasen, T., Dreibelbis, R., Saboori, S., Greene, L.E., Brumback, B., Muga, R., Rheingans, R., 2014. The impact of a school-based water supply and treatment, hygiene, and sanitation programme on pupil diarrhoea: a cluster-randomized trial. Epidemiology & Infection, 142(2), 340–351.

Groce, N., Bailey, N., Lang, R., Trani, J.F., Kett, M., 2011. Water and sanitation issues for persons with disabilities in low-and middle-income countries: a literature review and discussion of implications for global health and international development. J. Water Health 9 (4), 617–627.

Jalan, J., Somanathan, E., 2008. The importance of being informed: Experimental evidence on demand for environmental quality. J. Dev. Econ. 87, 14–28.

Mazengia, M.S., Chidavaenzi, M., Bradley, M., 2002. Effective and culturally acceptable water storage in Zimbabwe: maintaining the quality of water abstracted from upgraded family wells. J. Environ. Health 64, 15–18.

Momba, M.N.B., Kaleni, P., 2002. Regrowth and survival of indicator micro-organisms on the surfaces of household containers used for the storage of drinking water in rural communities of South Africa. Water Res. 36, 3023–3028.

Momba, M.N.B., Notshe, T.L., 2003. The effect of long storage in household containers on the microbiological quality of drinking water in rural communities of South Africa. J. Water Supply: Res. Technol. - AQUA 52(1), 67–77.

Momba, M.N.B., Malakate, V.K., Theron, J., 2006. Abundance of pathogenic Escherichia coli, Salmonella typhimurium and Vibrio cholerae in Nkonkobe drinking water sources. J. Water Health 04, 289–296.

Momba, M.N.B., Abongo’o, B.O., Mwambakana, J.N., 2008. Prevalence of enterohaemorrhagic Escherichia coli O157:H7 in drinking water and its predicted impact on diarrhoeic HIV/AIDS patients in the Amathole District, Eastern Cape Province, South Africa. Water SA 34, 365–372.

Peter, G., 2010. Impact of rural water projects on hygienic behaviour in Swaziland. Phys. Chem. Earth 35, 772–779.

Phaswana-Mafuya, N., 2006a. Health aspects of sanitation among Eastern Cape (EC) rural communities, South Africa. Curationis 29(2), 41–7.

Phaswana-Mafuya, N., 2006b. Hygiene status of rural communities in the Eastern Cape of South Africa. Int. J. Environ. Health Res. 16(4), 289–303.

Pond, K., Rueedi, J., Pedley, S., 2004. Microrisk: Pathogens in drinking water sources, Robens Centre for Public and Environmental health, University of Surrey. UK.

Quick, R., Venczel, L., Mintz, E., Soleto, L., Aparicio, J., Gironaz, M., Hutwagner, L., Greene, K., Bopp, C., Maloney, K., Chavez, D., Sobsey, M., Tauxe, R., 1999. Diarrhea prevention in Bolivia through point-of-use disinfection and safe storage: a promising new strategy. Epi Infect; 122: 83–90.

Statistics South Africa (StatsSA), 2011. Statistics South Africa, Millennium Development Goals, country report 2013. http://www.statssa.gov.za/MDG/MDGR_2013pdf. Accessed 24/06/2015.

Statistics South Africa, 2013. Millennium Development Goals, country report 2013. http://www.statssa.gov.za/MDG/MDGR_2013pdf. Accessed 24/06/2015.

Tshibangu, N.N., 1987. Water supplies and sanitary facilities in rural Transkei. S. Afr. Med. J. 17(6), 368–369.

United Nations, 2015. “Transforming our World: The 2030 Agenda for Sustainable Development”.

WHO/UNICEF Joint Monitoring Programme (JMP) for Water Supply and Sanitation, 2015. Progress on Sanitation and Drinking Water – 2015 Update and MDG Assessment. UNICEF and World Health Organization 2015, Geneva.

WHO/UNICEF, 2010. Progress on Sanitation and Drinking Water. WHO Press, Geneva, Switzerland.

WHO/UNICEF, 2013. Progress on Sanitation and Drinking-Water. 2013 Update. World Healzh Organization, Geneva.

